# Self-Care Confidence, Professional Support and Health Literacy in the UK: Findings from the Living Self-Care Survey Study

**DOI:** 10.1101/2025.06.01.25328745

**Authors:** Peter Smith, Sami Altalib, Mahmoud Al Ammouri, Austen El-Osta

**Author notes:** Corresponding author, (PS). Author Contributors: All authors provided substantial contributions to the conception (PS, AEO), design (AEO, PS, SA, MA), acquisition (SA, AEO) and interpretation (PS, SA, MA, DM, RB, AEO) of study data and approved the final version of the paper. AEO took the lead in planning the study with support from co-authors. SA and MA carried out the data analysis with support from AEO. AEO is the guarantor.

## Abstract

**Background:** Self-care is recognized as a cornerstone of modern health and care. The interface between health and care professionals (HCPs) and the public is pivotal in promoting self-care behaviours, yet barriers such as health literacy deficits, professional constraints, misconceptions and disparities in self-care confidence persist.

**Objective:** This study aimed to explore how self-care confidence, professional support and health literacy interact to influence self-care behaviours among UK adults, using a validated survey instrument to identify key disparities and enablers.

**Methods:** A cross-sectional online survey was used to collect data from 3,255 UK adults including health and care professionals between January and September 2024. It explored demographic characteristics, self-care knowledge, health literacy, engagement with digital health resources and professional encouragement. Data were analysed using descriptive statistics and ordinal logistic regression.

**Results:** Most respondents (88.5%) reported confidence in maintaining a healthy lifestyle; only 62.0% felt confident managing common illnesses. Healthcare guidance accessibility was perceived as easy (77.4%) but not evaluating treatment options (51.4%) or mental health information accessibility (43.6%). HCPs overwhelmingly endorsed the importance of self-care (94.7%) but barriers included reluctance of patients/clients to engage or take responsibility (64.8%), understanding (59.0%), time constraints (42.7%) and health literacy challenges (45.8%). 68.6% found screening information accessible but 28.1% struggled with interpretation. Regression analyses revealed that older adults (65+) were significantly less confident in self-care with professional guidance (aOR=0.50, p=0.001), whereas males (aOR=1.41, p<0.001) and Black/Asian British individuals (aORs=2.31, 1.98; p<0.001) reported higher confidence levels.

**Conclusion:** This study highlights the complex relationship between professional guidance, self-care confidence and health literacy. While most individuals value and engage in self-care, critical disparities persist, particularly in health literacy and access to digital resources. Targeted interventions and strengthening the public-healthcare professional dialogue and interface will be crucial in advancing self-care as a sustainable pillar of healthcare policy and practice.

## Introduction

Self-care is increasingly recognized as a critical component of healthcare systems worldwide, particularly as governments and public health organizations strive to improve population health outcomes while reducing healthcare burdens. The World Health Organization (WHO) defines self-care as “the ability of individuals, families and communities to promote health, prevent disease, maintain health and cope with illness and disability with or without the support of a healthcare provider” (1). Despite its recognized benefits, self-care implementation remains inconsistent across populations due to variations in confidence, knowledge, access to reliable health information and engagement with health and care professionals (HCPs).

The role of HCPs in promoting the adoption of heath-seeking self-care behaviours and evidence-based lifestyle medicine interventions is pivotal (2–4). Evidence suggests that professional guidance can significantly enhance self-care confidence (5), yet the extent to which individuals feel supported in this regard varies widely (3). While some populations may readily adopt self-care practices with minimal professional intervention, others require more structured guidance to navigate health decisions effectively. The extent of encouragement from HCPs, the perceived accessibility of healthcare services and the clarity of self-care information all influence the degree to which individuals can manage their own health. Professional perceptions of patient readiness for self-care and system-level constraints such as time pressures, resource limitations and varying levels of health literacy also affect how HCPs engage in promoting self-care (2,3).

Health literacy in particular has been identified as a major determinant of self-care efficacy(4)(6). Low health literacy is associated with poorer health outcomes, increased hospitalizations and reduced engagement in preventive healthcare behaviours. Digital health resources have emerged as a promising solution to bridge literacy gaps, but disparities remain in individuals’ ability to evaluate and apply online health information effectively. Given these challenges, there is a clear and urgent need to better understand the dynamic interface between HCPs and the public to identify both the enablers and barriers that influence effective self-care promotion.

Although a number of seminal studies have explored aspects of self-care (5)(7–9), there remains a significant gap in the availability of a consistent, validated set of survey questions that can be easily repeated over time. This limits our ability to track changes in knowledge, attitudes and behaviours related to self-care and hinders longitudinal analyses that could inform policy and practice. The Living Self-Care Survey Study was developed to address these limitations by providing a reproducible survey instrument that can be used in future research to track trends and inform self-care policy and practice.

The aim of this study was to explore self-care confidence, the role of HPCs in supporting self-care and the health literacy landscape among UK adults. Additionally, it sought to validate a reproducible survey instrument to support future research and policy development in this field. Specifically, it examined how health and care professionals influence self-care behaviours, the barriers they perceive and demographic disparities in self-care confidence. We also assessed the impact of health literacy on individuals’ ability to evaluate and apply health information and investigates whether digital health resources enhance or hinder self-care.

## Methods

### Study design

This study employed a cross-sectional online survey-based design to assess the interplay between self-care confidence, professional guidance and health literacy among community-dwelling adults in the UK. The study adhered to the Checklist for Reporting Results of Internet E-Surveys (CHERRIES) to ensure transparency, validity and reproducibility (10).

### Survey development

The survey was developed through a rigorous process involving literature review, adaptation of validated self-care assessment tools and expert consultation. Key sources used in survey development included The Department of Health’s Public Attitudes to Self-Care Baseline Survey (2005) that provided extensive data on public perceptions and behaviours (9), Elliott et al.’s Symptom Iceberg Study guided questions on symptom self-management and healthcare-seeking behaviour (8, 11), Smith et al.’s COVID-19 Self-Care Attitudes Study (2023) informed assessment of health and care professionals’ perspectives (12). The HLS19-Q12 (European Health Literacy Questionnaire)) provided a validated framework for measuring health literacy levels across demographic groups (9)(13). Additional questions on NHS-recommended health parameters for alcohol intake and exercise levels were included to allow a comparison to findings from 2005 surveys on the same (14, 15).

### Electronic survey

The survey was structured into six thematic blocks to collect data on (i) demographic characteristics, (ii) health and wellbeing, (iii) self-care knowledge and practices, (iv) barriers to self-care, (v) management of common symptoms and conditions, and (vi) questions specifically for respondents who are health and care professionals. Adaptive questioning was employed to optimize usability and engagement and ensure that only relevant questions were displayed based on prior responses. The order of the questions was fixed, and no randomisation or alteration was employed. The survey was uploaded in Qualtrics XM, before testing for usability and technical functionality. The questionnaire was piloted with six departmental colleagues and 15 participants, representing diverse demographics. Brief personal interviews and quantitative analyses were employed to assess question comprehension and psychometric properties. Based on pilot results and expert review, necessary modifications were made to enhance the survey’s validity and reliability. The synthesised Living Self-Care Survey can be accessed in S1 File.

### Participants

A total of 3,255 community-dwelling UK adults completed the survey. To ensure diversity, recruitment targeted both HCPs and the public. Demographic variables recorded included age, gender, ethnicity, employment type, highest education level, disability status and UK region of residence.

### Data Collection

The link to the open online survey was published and available on the Imperial College Qualtrics platform between 1 January and 30 September 2024 (9 months). The survey consisted of 83 questions displayed across 15 screens and was accessible on both personal computers and smartphones. To prevent participants from completing the survey more than once, Qualtrics XM places a browser cookie upon response submission, barring repeat attempts. The voluntary survey could be accessed by anyone with a link. Potentially eligible participants received an invitation email from the study team. The Self Care Forum also disseminated the email and link to a network of health and social care professionals.

Study information including the Participant Information Sheet (PIS) and link to the survey was also disseminated on various social media channels including X and LinkedIn. The researchers’ personal and professional networks were also mobilised to respond and further disseminate the survey among potentially eligible participants. The PIS included information regarding the study’s aims, the protection of participants’ personal data, the length of time of the survey, their right to withdraw from the study at any time, which data were stored, where and for how long, who the investigator was and survey length. Participants were informed that this was a voluntary survey without any monetary incentives but offering the possibility to access the findings at a later stage whilst underlying the potential collective benefits of taking part in terms of helping advance knowledge in this area and the formulation of future policies. The data collected were stored on the Imperial College London secure database and only the team researchers could access the survey results. All responses were pseudo-anonymised to ensure confidentiality by assigning each respondent a unique study ID. Only the participants’ demographic data (age in years, gender, ethnicity, employment type, highest education level and region of residence) were recorded. Respondents were able to refrain from providing an answer by selecting ’ prefer not to say’. Such answers were treated as missing data in all the analyses and complete case analysis was done (listwise exclusion). Due to the small number of missingness (<1.5%), the data were not imputed (16, 17).

### Data analysis

Quantitative data were collected using an eSurvey questionnaire administered on Qualtrics. Survey responses were summarised using frequencies and percentages. Ordinal logistic regression analyses were conducted to examine the relationship between demographic factors and self-care outcomes. The analyses were performed using both unadjusted and adjusted models. The adjusted logistic regression models controlled for age, gender, ethnicity, highest level of education, employment status, disability, long-term conditions, healthcare professional status and residency. For each outcome, odds ratios (OR) and adjusted odds ratios (aOR) with 95% confidence intervals (CI) were calculated. Statistical significance was set at p<0.05. The reference categories for categorical variables were consistently defined across analyses (e.g., age 18-24, female gender, White ethnicity). All analyses were performed using STATA, version 17 (StataCorp LP, College Station, TX, USA).

### Ethics

The study was reviewed and given ethical approval by Imperial College Research Ethics Committee (ICREC #6979141). Participants consented to take part in the survey by completing relevant tick box consent items at the start of the survey.

### Patient and Public Involvement

No patient was involved.

## Results

### Demographic profile of respondents

The survey included 3,255 respondents; the majority were aged between 25-34 years (22.9%) and 35-44 years (22.4%). Participant characteristics are shown in **Table 1**. Gender distribution was approximately balanced, with 52.1% identifying as female and 46.9% as male. The ethnic composition was predominantly White (82.4%), with smaller representations from Asian/Asian British (6.5%), Black/Black British (4.7%), Mixed/Multiple ethnic groups (3.8%) and other backgrounds (1.7%). Most participants resided in England (85.9%), held a university degree (45.6%), were employed full-time (48.9%) and 12.8% identified as having a disability.

**Table 1:**
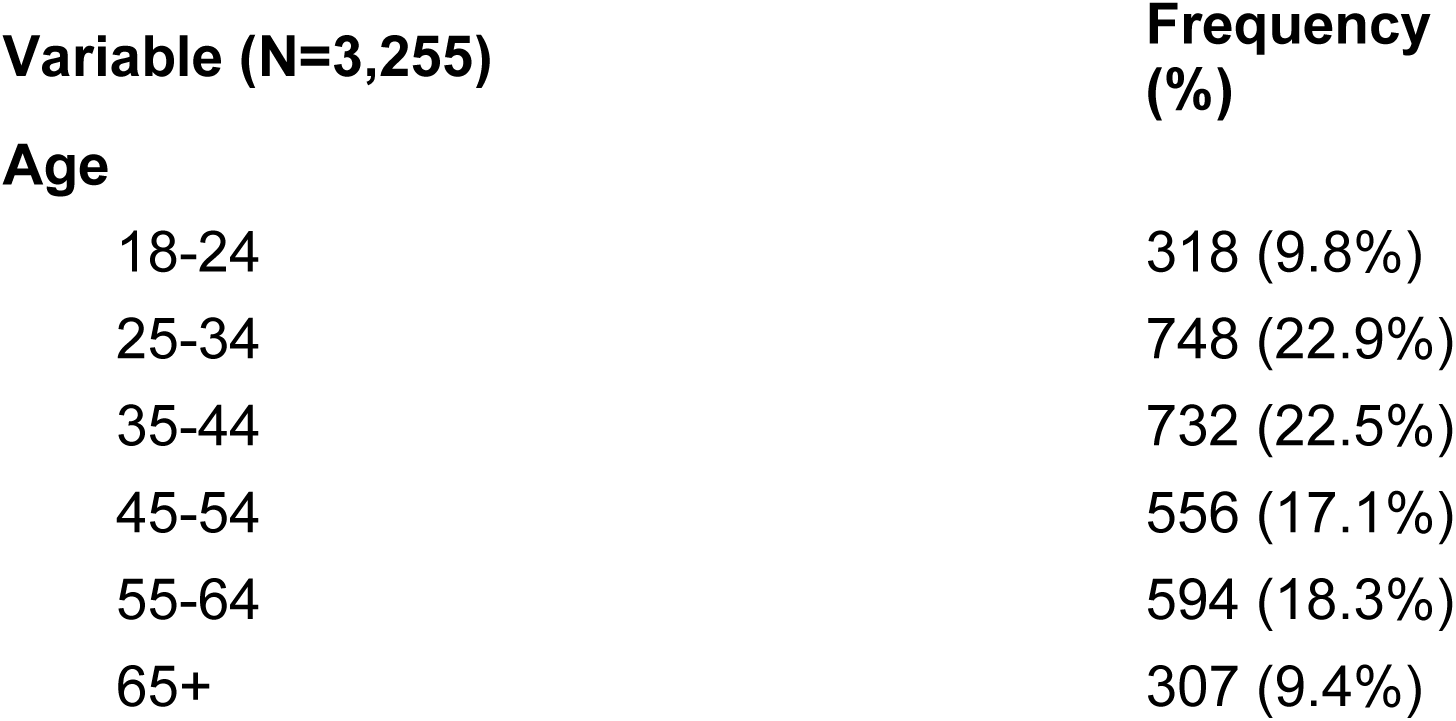
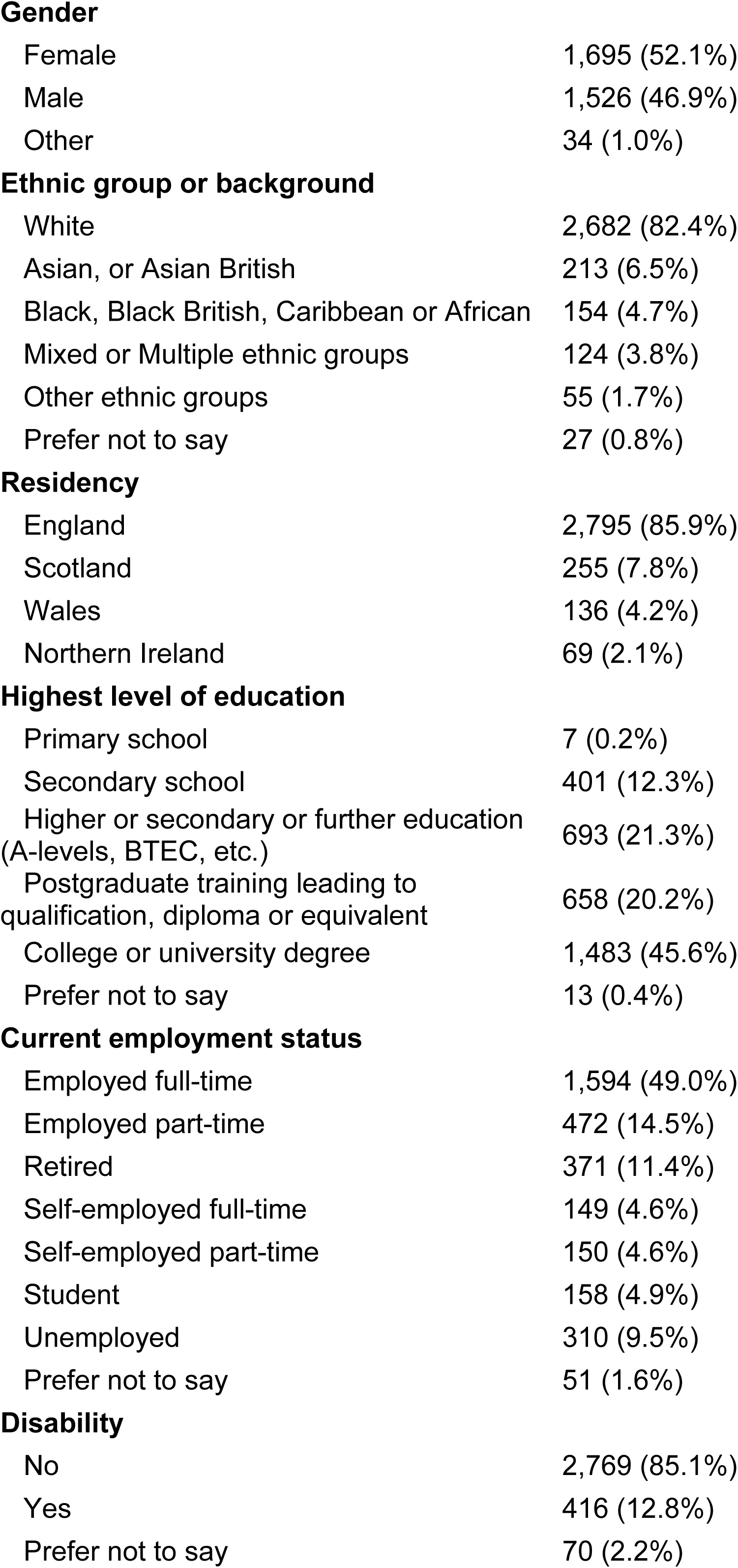
Participant characteristics.

| Variable (N=3,255) | Frequency (%) |
| --- | --- |
| **Age** |  |
| 18-24 | 318 (9.8%) |
| 25-34 | 748 (22.9%) |
| 35-44 | 732 (22.5%) |
| 45-54 | 556 (17.1%) |
| 55-64 | 594 (18.3%) |
| 65+ | 307 (9.4%) |
| **Gender** |  |
| Female | 1,695 (52.1%) |
| Male | 1,526 (46.9%) |
| Other | 34 (1.0%) |
| **Ethnic group or background** |  |
| White | 2,682 (82.4%) |
| Asian, or Asian British | 213 (6.5%) |
| Black, Black British, Caribbean or African | 154 (4.7%) |
| Mixed or Multiple ethnic groups | 124 (3.8%) |
| Other ethnic groups | 55 (1.7%) |
| Prefer not to say | 27 (0.8%) |
| **Residency** |  |
| England | 2,795 (85.9%) |
| Scotland | 255 (7.8%) |
| Wales | 136 (4.2%) |
| Northern Ireland | 69 (2.1%) |
| **Highest level of education** |  |
| Primary school | 7 (0.2%) |
| Secondary school | 401 (12.3%) |
| Higher or secondary or further education<br>(A-levels, BTEC, etc.) | 693 (21.3%) |
| Postgraduate training leading to<br>qualification, diploma or equivalent | 658 (20.2%) |
| College or university degree | 1,483 (45.6%) |
| Prefer not to say | 13 (0.4%) |
| **Current employment status** |  |
| Employed full-time | 1,594 (49.0%) |
| Employed part-time | 472 (14.5%) |
| Retired | 371 (11.4%) |
| Self-employed full-time | 149 (4.6%) |
| Self-employed part-time | 150 (4.6%) |
| Student | 158 (4.9%) |
| Unemployed | 310 (9.5%) |
| Prefer not to say | 51 (1.6%) |
| **Disability** |  |
| No | 2,769 (85.1%) |
| Yes | 416 (12.8%) |
| Prefer not to say | 70 (2.2%) |

### Main survey findings

The results of the main survey are shown in S2 File.

### Accessing professional support

Most participants found accessing professional help straightforward, with 77.4% reporting it was “very easy” or “easy,” similar to understanding emergency medical information (77.7%). Only 50.5% found mental health information accessible with the remaining 43.6% reporting difficulties. Access to health screening information was also split, with 68.6% reporting ease and 28.1% indicating difficulties. Evaluating treatment options was challenging for 51.4%.

### Perceived importance of self-care

Of the 227 HCPs surveyed, 76.2% considered self-care “very important” and 94.7% considered it important or very important.

### Professional view of barriers to self-care

HCPs identified multiple barriers that hinder patients from effectively engaging in self-care (S1 Table). The most frequently cited obstacles were patients’ reluctance to take responsibility for their own care (64.8%), followed by a poor understanding of self-care (59.0%), highlighting the need for enhanced educational initiatives and motivational strategies to encourage patient participation in self-care. Structural challenges, including health inequalities (52.0%) and time constraints (42.7%), were also significant barriers, reflecting systemic issues that limit patient autonomy. Additionally, limited access to digital resources and low digital skills (26.4%) emerged as obstacles, potentially excluding certain populations from accessing self-care opportunities available through online platforms. Dependency on HCPs (31.7%) and inconsistent messaging among providers (28.2%) further complicated patient engagement, highlighting the need for standardized self-care guidance. Health literacy deficits (45.8%) and communication barriers (32.6%) were also prominent concerns, reinforcing the importance of providing clear, accessible health information to patients. Although fewer respondents cited a lack of appropriate resources (15.9%) and professional misunderstandings of self-care (12.8%) as barriers, these factors still point to gaps in both patient-facing and provider-level education.

### Impact of NHS Guidelines and Professional Support

Despite high confidence in knowledge and understanding to lead a healthy lifestyle (88.5% very confident or fairly confident), significant knowledge gaps persisted in key health parameters, including physical activity and dietary recommendations. When asked about NHS guidelines, 54.8% correctly identified the recommended weekly alcohol intake limits for males, and only 23.3% were aware of the Chief Medical Officer’s advice for all to engage in 150 minutes of moderate exercise per week. Both public and professional groups demonstrated similar levels of knowledge.

### Health Literacy – HLS19-Q12

Health literacy findings revealed disparities in locating, understanding and applying health information. Table 2 includes the combined percentage of those reporting that items were ‘difficult’ or ‘very difficult’ to achieve, allowing comparisons with 17 Eurasian countries that have previously used the HLS19-Q12 survey. Only 21.1% reported difficulties in accessing professional healthcare, whereas 20.6% found it difficult to understand emergency medical guidance. More than half (51.4%) struggled to evaluate treatment options and 43.6% faced difficulties finding mental health resources.

**Table 2:** Health Literacy country comparison. Scores ranked by the responses to the HLS19-Q12 from the Living Self-Care Survey: “On a scale from ‘very easy’ to ‘very difficult’, how easy would you say it is to…”

| **Respondents finding it ‘difficult’ or ‘very difficult’ to:** | **%** | **Rank\***<br>**/18** |
| --- | --- | --- |
| Judge the advantages & disadvantages of different treatment options | 51.4 | 16 |
| Decide how you can protect yourself from illness using information from the mass media | 49.9 | 15 |
| Find information on how to handle mental health problems | 43.6 | 15 |
| Make decisions to improve your health & wellbeing | 26.9 | 10 |
| Understand information about what to do in a medical emergency | 20.6 | 7 |
| Judge how your housing conditions may affect your health & wellbeing | 33.7 | 16 |
| Judge if information on unhealthy habits, such as smoking, low physical activity or drinking too much alcohol, are reliable | 20.7 | 12 |
| Understand information about recommended health screenings or examinations? | 28.1 | 15 |
| Understand advice concerning your health from family or friends | 24.3 | 17 |
| Find out where to get professional help when you are ill | 21.1 | 14 |
| Find information on healthy lifestyles such as physical exercise, healthy food or nutrition | 12.9 | 14 |
| Act on advice from your doctor or pharmacist | 13.0 | 15 |
| * UK Rank Compared to 17 other Eurasian countries: Austria, Belgium, Bulgaria, Czech Republic, Denmark, France, Germany, Hungary, Ireland, Israel, Italy, Norway, Portugal, Russian Federation, Slovakia, Slovenia, Switzerland. |  |  |

Nearly a third (28.1%) had trouble understanding information on health screenings, which could impact preventive care engagement. Decision-making based on mass media health information was a notable challenge, with 49.9% reporting difficulty in using such sources for health protection. In contrast, only12.9% found information on physical activity and nutrition difficult to find. Understanding the impact of housing on health posed challenges for 33.7% of respondents, suggesting gaps in environmental health literacy. Although most respondents were confident following medical advice, 13% found it difficult or very difficult, indicating barriers beyond comprehension, such as trust, accessibility, or socioeconomic constraints. These findings emphasize the need for targeted interventions to enhance health literacy, particularly in evaluating treatment options, making informed health decisions using digital media and improving mental health awareness. (S2 Table).

## Inferential findings

### Confidence in self-care with professional guidance

The ordinal logistic regression analysis assessed demographic factors influencing confidence in caring for one’s own health with guidance and support from health and care professionals (Table 3, Fig 1). Age and ethnicity significantly predicted confidence in self-care with NHS guidance. Older adults (aged 65+) had half the odds than younger participants (18–24) to believe they would be more confident with NHS support (aOR = 0.50, p = 0.001). Males (aOR = 1.41, p < 0.001) and Black and Asian British individuals (aORs = 2.31, 1.98; p < 0.001) had significantly higher odds in reporting increased confidence with professional guidance.

**Fig 1:**
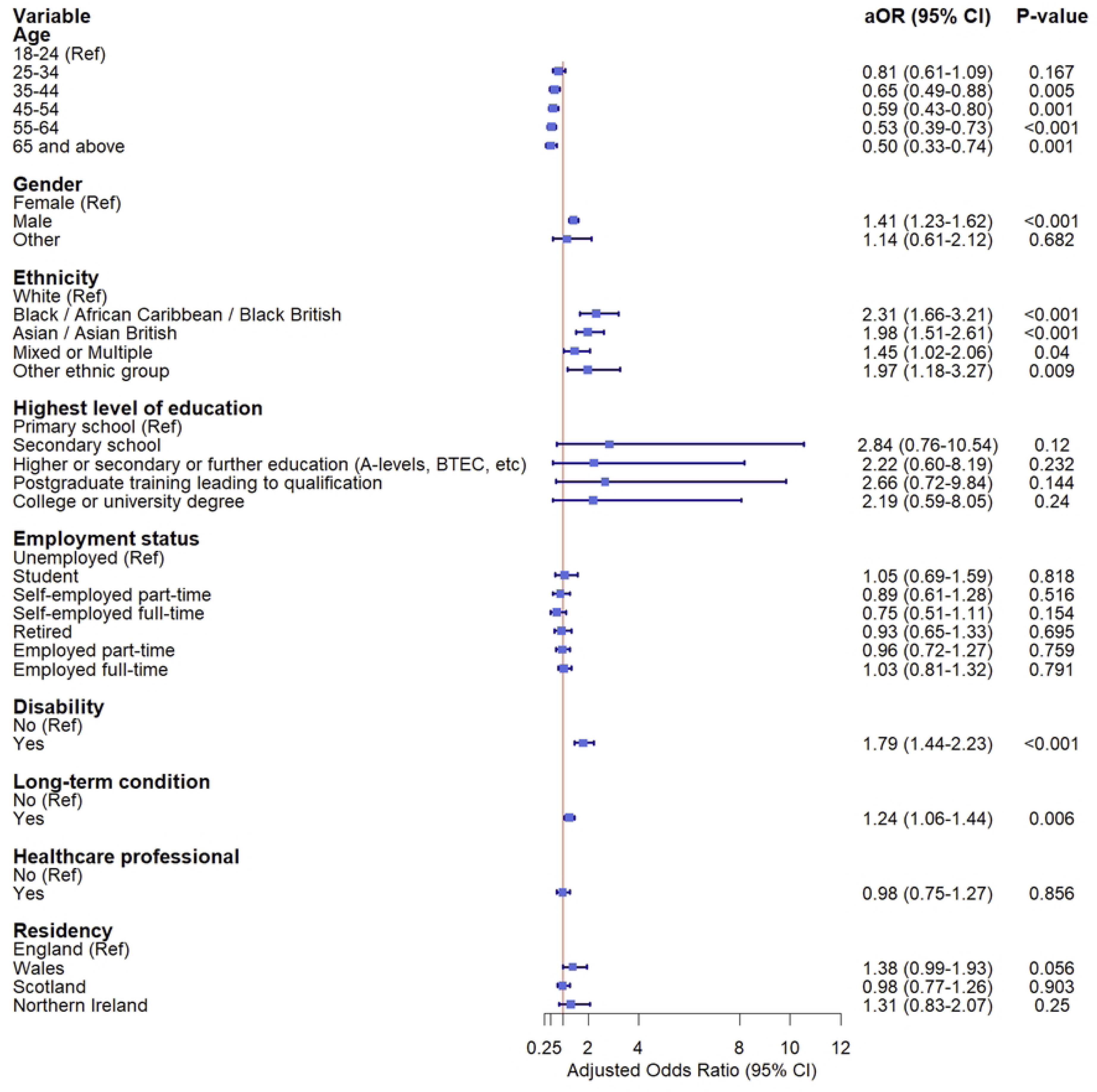
Factors affecting confidence caring for own health with guidance and support from HCP. Forest plot showing the odds ratios and confidence intervals of factors associated with being more confident about caring for own health with guidance & support from an HCP.

**Table 3:** Ordinal logistic regression analysis of demographic factors predicting confident about caring for my own health if I had guidance & support from HCP.

| Variable | being more confident about caring for own health with guidance & support from an NHS professional |  |  |  |
| --- | --- | --- | --- | --- |
|  | Unadjusted |  | Adjusted |  |
|  | OR (95% CI) | P-value | aOR (95%) | P- |
| **Age** |  |  |  |  |
| 18-24 | Ref |  | Ref |  |
| 25-34 | 0.81 (0.64 - 1.04) | 0.095 | 0.81 (0.61 - 1.09) | 0.167 |
| 35-44 | 0.62 (0.49 - 0.79) | **<0.001** | 0.65 (0.49 - 0.88) | **0.005** |
| 45-54 | 0.61 (0.48 - 0.79) | **<0.001** | 0.59 (0.43 - 0.80) | **0.001** |
| 55-64 | 0.51 (0.40 - 0.65) | <0.001 | 0.53 (0.39 - 0.73) | <0.001 |
| 65 and above | 0.48 (0.36 - 0.64) | <0.001 | 0.50 (0.33 - 0.74) | 0.001 |
| **Gender** |  |  |  |  |
| Female | Ref |  | Ref |  |
| Male | 1.39 (1.22 - 1.57) | <0.001 | 1.41 (1.23 - 1.62) | <0.001 |
| Other | 1.45 (0.79 - 2.66) | 0.236 | 1.14 (0.61 - 2.12) | 0.682 |
| **Ethnicity** |  |  |  |  |
| White | Ref |  | Ref |  |
| Black / African Caribbean / Black British | 2.20 (1.62 - 2.98) | <0.001 | 2.31 (1.66 - 3.21) | <0.001 |
| Asian / Asian British | 2.04 (1.59 - 2.63) | <0.001 | 1.98 (1.51 - 2.61) | <0.001 |
| Mixed or Multiple | 1.60 (1.15 - 2.22) | 0.006 | 1.45 (1.02 - 2.06) | 0.040 |
| Other ethnic group | 1.76 (1.08 - 2.87) | 0.023 | 1.97 (1.18 - 3.27) | 0.009 |
| **Highest level of education** |  |  |  |  |
| Primary school | Ref |  | Ref |  |
| Secondary school | 1.35 (0.34 - 5.32) | 0.669 | 2.84 (0.76 - 10.54) | 0.120 |
| Higher or secondary or further education (A-levels, BTEC, etc) | 1.23 (0.31 - 4.84) | 0.763 | 2.22 (0.60 - 8.19) | 0.232 |
| Postgraduate training leading to qualification | 1.45 (0.37 - 5.70) | 0.593 | 2.66 (0.72 - 9.84) | 0.144 |
| College or university degree | 1.24 (0.32 - 4.86) | 0.753 | 2.19 (0.59 - 8.05) | 0.240 |
| **Employment status** |  |  |  |  |
| Unemployed | Ref |  | Ref |  |
| Student | 1.39 (0.98 - 1.97) | 0.063 | 1.05 (0.69 - 1.59) | 0.818 |
| Self-employed part-time | 0.66 (0.47 - 0.93) | 0.018 | 0.89 (0.61 - 1.28) | 0.516 |
| Self-employed full-time | 0.59 (0.41 - 0.84) | 0.004 | 0.75 (0.51 - 1.11) | 0.154 |
| Retired | 0.66 (0.50 - 0.87) | 0.003 | 0.93 (0.65 - 1.33) | 0.695 |
| Employed part-time | 0.75 (0.58 - 0.97) | 0.030 | 0.96 (0.72 - 1.27) | 0.759 |
| Employed full-time | 0.92 (0.74 - 1.15) | 0.455 | 1.03 (0.81 - 1.32) | 0.791 |
| **Disability** |  |  |  |  |
| No | Ref |  | Ref |  |
| Yes | 1.73 (1.43 - 2.09) | <0.001 | 1.79 (1.44 - 2.23) | <0.001 |
| **Long-term condition** |  |  |  |  |
| No | Ref |  | Ref |  |
| Yes | 1.16 (1.02 - 1.32) | **0.022** | 1.24 (1.06 - 1.44) | **0.006** |
| **Healthcare professional** |  |  |  |  |
| No | Ref |  | Ref |  |
| Yes | 1.08 (0.84 - 1.38) | 0.561 | 0.98 (0.75 - 1.27) | 0.856 |
| **Residency** |  |  |  |  |
| England | Ref |  | Ref |  |
| Wales | 1.32 (0.96 - 1.81) | 0.090 | 1.38 (0.99 - 1.93) | 0.056 |
| Scotland | 0.91 (0.72 - 1.15) | 0.444 | 0.98 (0.77 - 1.26) | 0.903 |
| Northern Ireland | 1.24 (0.81 - 1.91) | 0.324 | 1.31 (0.83 - 2.07) | 0.250 |

*Adjusted for age, gender, ethnicity, highest level of education, employment status, disability, long-term conditions, being a healthcare professional or not and residency. (Outcome: Strongly disagree, Disagree, Neither agree nor disagree, Agree or Strongly agree)*

### Desire for more health responsibility

The ordinal logistic regression analysis (Table 4, Fig 2) examined demographic factors associated with the agreement with the statement “I don’t want any more responsibility over my health”. Older adults had lower odds in desiring additional health responsibility, with the odds of agreeing diminishing by over half in participants aged 65+ (aOR=0.42, p<0.001). Black and Asian British respondents were also less inclined to want more responsibility compared to White participants (aORs=0.69, 0.73; p<0.05).

**Fig 2:**
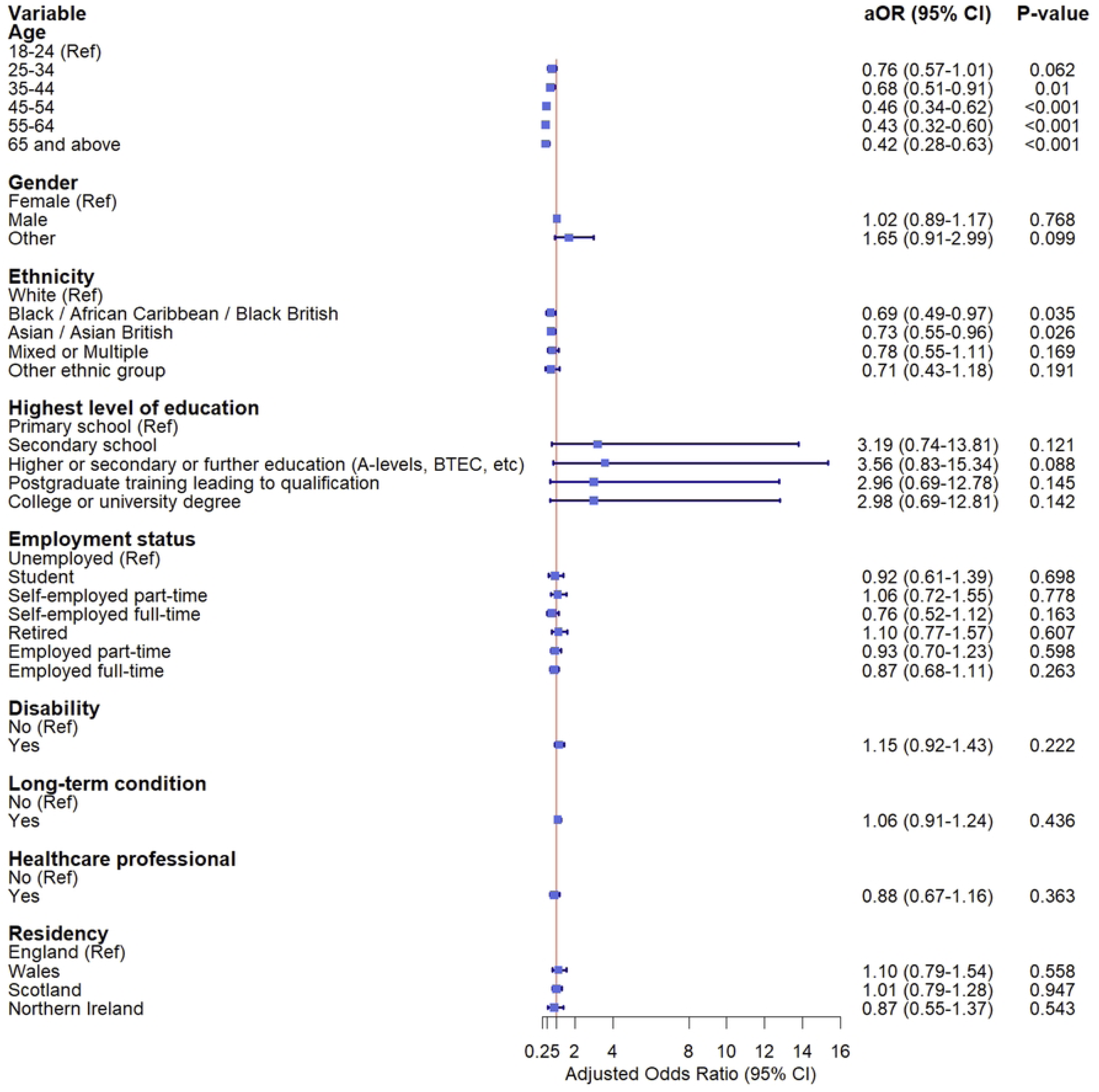
Factors associated with not wanting more responsibility over own health. Forest plot showing the odds ratios and confidence intervals of factors associated with not wanting more responsibility over own health

**Table 4:** Ordinal logistic regression analysis of demographic factors predicting I don’t want any more responsibility over my health.

| Variable | Not wanting more responsibility over own health |  |
| --- | --- | --- |
|  | Unadjusted | Adjusted |

|  | OR (95% CI) | P-value | aOR (95% CI) | P-value |
| --- | --- | --- | --- | --- |
| **Age** |  |  |  |  |
| 18-24 | Ref |  | Ref |  |
| 25-34 | 0.77 (0.61 - 0.98) | **0.037** | 0.76 (0.57 - 1.01) | 0.062 |
| 35-44 | 0.68 (0.53 - 0.86) | **0.001** | 0.68 (0.51 - 0.91) | **0.010** |
| 45-54 | 0.50 (0.39 - 0.65) | **<0.001** | 0.46 (0.34 - 0.62) | **<0.001** |
| 55-64 | 0.50 (0.39 - 0.65) | **<0.001** | 0.43 (0.32 - 0.60) | **<0.001** |
| 65 and above | 0.56 (0.42 - 0.74) | **<0.001** | 0.42 (0.28 - 0.63) | **<0.001** |
| **Gender** |  |  |  |  |
| Female | Ref |  | Ref |  |
| Male | 1.00 (0.89 - 1.14) | 0.948 | 1.02 (0.89 - 1.17) | 0.768 |
| Other | 1.74 (0.97 - 3.12) | 0.064 | 1.65 (0.91 - 2.99) | 0.099 |
| **Ethnicity** |  |  |  |  |
| White | Ref |  | Ref |  |
| Black / African Caribbean / Black British | 0.76 (0.55 - 1.04) | 0.084 | 0.69 (0.49 - 0.97) | **0.035** |
| Asian / Asian British | 0.88 (0.68 - 1.13) | 0.318 | 0.73 (0.55 - 0.96) | **0.026** |
| Mixed or Multiple | 0.96 (0.69 - 1.33) | 0.797 | 0.78 (0.55 - 1.11) | 0.169 |
| Other ethnic group | 0.84 (0.52 - 1.37) | 0.494 | 0.71 (0.43 - 1.18) | 0.191 |
| **Highest level of education** |  |  |  |  |
| Primary school | Ref |  | Ref |  |
| Secondary school | 2.86 (0.69 - 11.86) | 0.147 | 3.19 (0.74 - 13.81) | 0.121 |
| Higher or secondary or further education (A-levels, BTEC, etc) | 3.55 (0.86 - 14.63) | 0.080 | 3.56 (0.83 - 15.34) | 0.088 |
| Postgraduate training leading to qualification | 2.66 (0.65 - 10.98) | 0.175 | 2.96 (0.69 - 12.78) | 0.145 |
| College or university degree | 2.85 (0.69 - 11.70) | 0.146 | 2.98 (0.69 - 12.81) | 0.142 |
| **Employment status** |  |  |  |  |
| Unemployed | Ref |  | Ref |  |
| Student | 1.14 (0.80 - 1.61) | 0.467 | 0.92 (0.61 - 1.39) | 0.698 |
| Self-employed part-time | 0.80 (0.56 - 1.14) | 0.213 | 1.06 (0.72 - 1.55) | 0.778 |
| Self-employed full-time | 0.63 (0.44 - 0.89) | **0.010** | 0.76 (0.52 - 1.12) | 0.163 |
| Retired | 0.76 (0.58 - 1.00) | **0.047** | 1.10 (0.77 - 1.57) | 0.607 |
| Employed part-time | 0.78 (0.60 - 1.01) | 0.061 | 0.93 (0.70 - 1.23) | 0.598 |
| Employed full-time | 0.78 (0.62 - 0.97) | **0.025** | 0.87 (0.68 - 1.11) | 0.263 |
| **Disability** |  |  |  |  |
| No | Ref |  | Ref |  |
| Yes | 1.22 (1.01 - 1.48) | **0.043** | 1.15 (0.92 - 1.43) | 0.222 |
| **Long-term condition** |  |  |  |  |
| No | Ref |  | Ref |  |
| Yes | 1.01 (0.88 - 1.15) | 0.917 | 1.06 (0.91 - 1.24) | 0.436 |
| **Healthcare professional** |  |  |  |  |
| No | Ref |  | Ref |  |
| Yes | 0.89 (0.69 - 1.15) | 0.376 | 0.88 (0.67 - 1.16) | 0.363 |
| **Residency** |  |  |  |  |
| England | Ref |  | Ref |  |
| Wales | 1.11 (0.81 - 1.53) | 0.510 | 1.10 (0.79 - 1.54) | 0.558 |
| Scotland | 0.98 (0.78 - 1.23) | 0.393 | 1.01 (0.79 - 1.28) | 0.947 |
| Northern Ireland | 0.96 (0.62 - 1.47) | 0.139 | 0.87 (0.55 - 1.37) | 0.543 |

*Adjusted for age, gender, ethnicity, highest level of education, employment status, disability, long-term conditions, being a healthcare professional or not and residency. Outcome: Strongly disagree, Disagree, Neither agree nor disagree, Agree or Strongly agree)*

### Encouragement from health and care professionals

Encouragement to self-care varied significantly by demographics (Table 5, Fig 3). Black British and Asian British participants had higher odds of feeling encouraged by healthcare professionals (aORs=1.58 and 1.45, respectively; p<0.05). Males had higher odds of reporting encouragement compared to females (aOR=1.20, p=0.012), whereas older adults (aged 55–64) had lower odds than younger counterparts (aOR=0.68, p=0.019).

**Fig 3:**
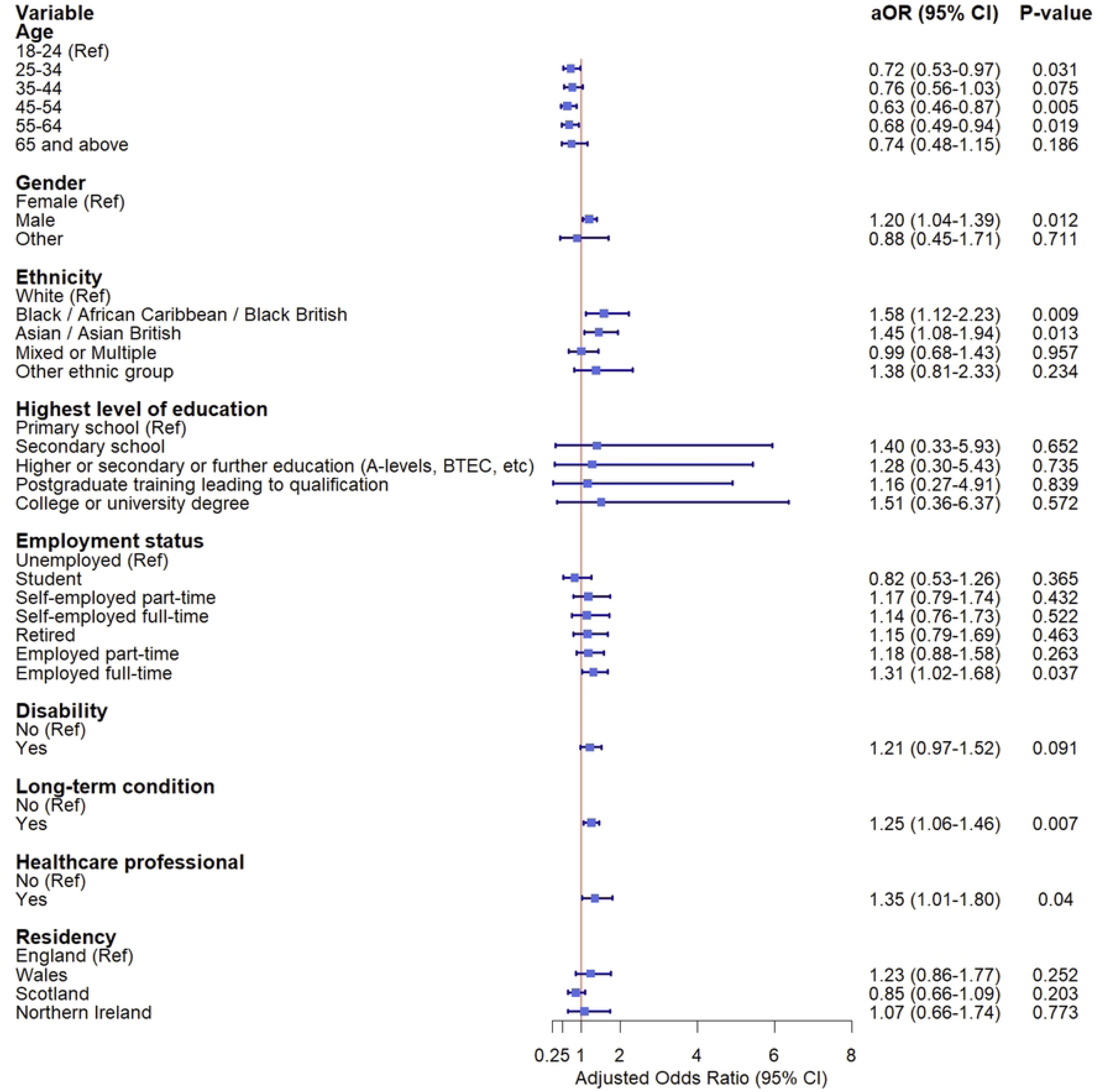
Encouragement by HCP to play more active role in staying healthy and treating common conditions. Forest plot showing the odds ratios and confidence intervals of factors associated with being encouraged by GP/nurse/pharmacist to play a more active role in staying healthy and treating common conditions by oneself

**Table 5:** Ordinal logistic regression analysis of demographic factors predicting I’m encouraged by my HCP to play a more active role in staying healthy & treating common conditions myself.

| Variable | Encouraged by GP/nurse/pharmacist to play more active role in staying healthy & treating common conditions by oneself |  |  |  |
| --- | --- | --- | --- | --- |
|  | Unadjusted<br>OR (95% CI) | P-value | Adjusted<br>aOR (95% CI) | P-value |
| **Age** |  |  |  |  |
| 18-24 | Ref |  | Ref |  |
| 25-34 | 0.84 (0.65 - 1.08) | 0.177 | 0.72 (0.53 - 0.97) | **0.031** |
| 35-44 | 0.87 (0.67 - 1.12) | 0.276 | 0.76 (0.56 - 1.03) | 0.075 |
| 45-54 | 0.72 (0.56 - 0.94) | **0.017** | 0.63 (0.46 - 0.87) | **0.005** |
| 55-64 | 0.74 (0.57 - 0.96) | **0.025** | 0.68 (0.49 - 0.94) | **0.019** |
| 65 and above | 0.81 (0.60 - 1.09) | 0.164 | 0.74 (0.48 - 1.15) | 0.186 |
| **Gender** |  |  |  |  |
| Female | Ref |  | Ref |  |
| Male | 1.23 (1.07 - 1.40) | **0.003** | 1.20 (1.04 - 1.39) | **0.012** |
| Other | 0.96 (0.50 - 1.82) | 0.890 | 0.88 (0.45 - 1.71) | 0.711 |
| **Ethnicity** |  |  |  |  |
| White | Ref |  | Ref |  |
| Black / African Caribbean / Black British | 1.62 (1.18 - 2.22) | **0.003** | 1.58 (1.12 - 2.23) | **0.009** |
| Asian / Asian British | 1.43 (1.09 - 1.86) | **0.009** | 1.45 (1.08 - 1.94) | **0.013** |
| Mixed or Multiple | 0.99 (0.70 - 1.42) | 0.982 | 0.99 (0.68 - 1.43) | 0.957 |
| Other ethnic group | 1.36 (0.82 - 2.24) | 0.234 | 1.38 (0.81 - 2.33) | 0.234 |
| **Highest level of education** |  |  |  |  |
| Primary school | Ref |  | Ref |  |
| Secondary school | 0.86 (0.21 - 3.55) | 0.837 | 1.40 (0.33 - 5.93) | 0.652 |
| Higher or secondary or further education (A-levels, BTEC, etc) | 0.84 (0.20 - 3.44) | 0.809 | 1.28 (0.30 - 5.43) | 0.735 |
| Postgraduate training leading to qualification | 0.84 (0.20 - 3.45) | 0.810 | 1.16 (0.27 - 4.91) | 0.839 |
| College or university degree | 1.03 (0.25 - 4.22) | 0.963 | 1.51 (0.36 - 6.37) | 0.572 |
| **Employment status** |  |  |  |  |
| Unemployed | Ref |  | Ref |  |
| Student | 1.11 (0.77 - 1.61) | 0.577 | 0.82 (0.53 - 1.26) | 0.365 |
| Self-employed part-time | 1.10 (0.76 - 1.59) | 0.627 | 1.17 (0.79 - 1.74) | 0.432 |
| Self-employed full-time | 0.96 (0.65 - 1.40) | 0.823 | 1.14 (0.76 - 1.73) | 0.522 |
| Retired | 1.16 (0.87 - 1.54) | 0.323 | 1.15 (0.79 - 1.69) | 0.463 |
| Employed part-time | 1.16 (0.88 - 1.52) | 0.291 | 1.18 (0.88 - 1.58) | 0.263 |
| Employed full-time | 1.30 (1.03 - 1.63) | **0.028** | 1.31 (1.02 - 1.68) | **0.037** |
| **Disability** |  |  |  |  |
| No | Ref |  | Ref |  |
| Yes | 1.18 (0.97 - 1.44) | 0.093 | 1.21 (0.97 - 1.52) | 0.091 |
| **Long-term condition** |  |  |  |  |
| No | Ref |  | Ref |  |
| Yes | 1.17 (1.02 - 1.34) | **0.025** | 1.25 (1.06 - 1.46) | **0.007** |
| **Healthcare professional** |  |  |  |  |
| No | Ref |  | Ref |  |
| Yes | 1.40 (1.07 - 1.83) | **0.013** | 1.35 (1.01 - 1.80) | **0.040** |
| **Residency** |  |  |  |  |
| England | Ref |  | Ref |  |
| Wales | 1.20 (0.85 - 1.69) | 0.304 | 1.23 (0.86 - 1.77) | 0.252 |
| Scotland | 0.87 (0.68 - 1.11) | 0.250 | 0.85 (0.66 - 1.09) | 0.203 |
| Northern Ireland | 1.04 (0.66 - 1.66) | 0.855 | 1.07 (0.66 - 1.74) | 0.773 |

*Adjusted for age, gender, ethnicity, highest level of education, employment status, disability, long-term conditions, being a healthcare professional or not and residency. (Outcome: Never, Sometimes, Usually or Always)*

## Discussion

### Summary of principal findings

To our knowledge, this is the first nationally representative UK study to examine the intersection of self-care confidence, professional guidance and health literacy. The study also helped introduce a reproducible survey instrument developed through expert consultation and piloted for usability, designed to support future longitudinal research into self-care attitudes and behaviours. The principal findings recapitulated below provide critical insights into how different population groups interact with healthcare professionals and self-care resources, revealing disparities in support, confidence and access to information.

#### Confidence in self-care and professional guidance

The high levels of confidence reported by participants in leading a healthy lifestyle (89%) contrast with their lower confidence in managing common illnesses (62%). This discrepancy aligns with prior research suggesting that while individuals may possess general health awareness, they often lack the necessary skills or assurance to manage acute or chronic conditions independently The significant association between professional guidance and increased self-care confidence underscores the indispensable role of HCPs in fostering self-efficacy in self-management However, our regression analysis indicates that older adults (65+) were significantly less likely to report enhanced confidence with professional guidance, which may reflect generational differences in health-seeking behaviours or digital health engagement.

#### Barriers to self-care engagement

Health and care professionals identified several barriers impeding patients’ engagement with self-care, including individual reluctance (65%), inadequate self-care understanding (60%) and health inequalities (52%). These findings resonate with studies emphasizing the critical role of health literacy in self-care adoption (18–20). Notably, health literacy deficits were perceived as a significant challenge by 46% of HCPs reinforcing the necessity for tailored educational interventions that improve patient comprehension and application of health information. Structural barriers such as time constraints (43%) and digital exclusion (26%) highlight systemic limitations that must be addressed to create equitable access to self-care resources.

#### Health literacy and digital health resources

Health literacy emerged as a pivotal determinant of self-care efficacy, with substantial proportions of respondents struggling to evaluate treatment options (51%) and access mental health information (44%). These findings align with previous studies demonstrating the association between low health literacy and poorer health outcomes (4)(21)(22). The challenges associated with assessing mass media health information (50%) further necessitate the critical appraisal of skills in an era dominated by digital health resources. While digital platforms offer vast opportunities for self-care empowerment, disparities in digital literacy and accessibility must be addressed to prevent exacerbation of health inequalities. Pertinently, our use of the HLS19-Q12 validated tool to measure health literacy allowed a comparison with 17 other countries. We found that UK respondents experience greater difficulty with health literacy than other Eurasian countries (8)(13).

#### Encouragement from health and care professionals

That encouragement from HCPs varied significantly across demographic groups, with Black and Asian British participants 1.6 more likely to report feeling supported in self-care suggests potential cultural or community-driven influences that may shape the reception and effectiveness of self-care messaging. Conversely, older adults (55–64) were less likely to feel encouraged by their HCPs, indicating possible gaps in tailored communication strategies. The role of personalized, culturally competent self-care guidance warrants further exploration to optimize engagement across diverse populations.

### Implications for policy and practice

To our knowledge, this is the largest and most comprehensive study globally to investigate the interplay between self-care confidence, professional support and health literacy in a general population sample. It offers important and actionable insights into how health systems, professionals and the public interact within the self-care landscape. The findings have several far-reaching implications for healthcare policy, system design, workforce development, and patient and public engagement strategies.

#### Institutionalising self-care as a core function of health systems

There is a compelling case to reframe self-care from being an optional, peripheral activity to a core, supported function of modern health systems. This study demonstrates that while individuals overwhelmingly express a willingness and perceived confidence in maintaining a healthy lifestyle, their capacity to self-manage common illnesses is significantly lower, indicating a systemic gap in self-care enablement. Health policy should respond by embedding self-care into national strategies, clinical guidelines, and population health frameworks. This includes the formal recognition of self-care as a care modality with structured referral pathways, care protocols, and outcome indicators.

#### Embedding self-care in professional education and training

The disconnect between public willingness to self-care and HCP-perceived barriers, including patient reluctance and low literacy, signals a need for enhanced professional development. Regulatory and educational bodies should integrate structured training on self-care facilitation, health coaching, and motivational interviewing into undergraduate and continuous professional development (CPD) curricula for all health and care professionals. This training should prioritise culturally responsive communication, shared decision-making, and the use of validated tools to assess readiness and capacity for self-care.

#### Addressing the health literacy divide as a matter of equity

The study’s health literacy findings highlight stark disparities in the ability to access, understand, evaluate and act on health information particularly around treatment options and mental health. These gaps should be recognised as a public health equity issue. National strategies should be developed to enhance health literacy at scale through public health campaigns, digital inclusion initiatives, and curriculum reform. Health literacy needs to be measured routinely, much like other health indicators, with efforts tailored to reach marginalised and digitally excluded communities.

#### Advancing digital health literacy through inclusive design

As digital platforms increasingly become the primary mode for accessing health information and services, this study’s evidence of digital exclusion must prompt urgent action. Policy and practice must ensure that digital health tools are not only available but also usable by people with varied literacy levels, disabilities, and socioeconomic backgrounds. Co-design with end-users, plain language content, and digital navigation support should become standard features of health technologies. Health systems should invest in digital health literacy as a foundational enabler of safe, effective, and equitable self-care.

#### Personalising self-care interventions through population segmentation

Given the variation in confidence, support, and desire for responsibility observed across age, gender, ethnicity, and disability status, self-care initiatives must be tailored to the needs and preferences of different groups. For example, older adults and those with long-term conditions may benefit from structured self-care support integrated into chronic disease management, whereas younger adults may respond better to digital tools and peer support networks. Population segmentation approaches, already common in marketing and behavioural science, should be applied to self-care to maximise reach, relevance, and effectiveness.

#### Enabling primary care teams to act as self-care enablers, not just gatekeepers

Primary care settings are well-positioned to operationalise self-care as a first-line intervention, but this will require shifts in incentives, roles, and workflows. This includes adjusting appointment lengths to allow for self-care conversations, using technology to deliver asynchronous follow-ups, and enabling multidisciplinary teams (e.g. pharmacists, link workers, health coaches) to support ongoing self-care outside the consulting room. Payment models should reward prevention and self-care support activities, and outcome frameworks should capture patient activation, confidence, and self-efficacy.

#### Monitoring and evaluating self-care readiness and impact

This study also introduces a validated and reproducible tool to assess self-care confidence and health literacy, enabling regular tracking over time. National surveys or health service datasets could incorporate these items to monitor trends, identify high-need populations, and evaluate the impact of policies and interventions. The development of a national self-care index or dashboard could provide transparency, accountability, and continuous improvement in this area.

#### Creating a supportive policy ecosystem through multi-sectoral collaboration

Lastly, realising the potential of self-care requires action beyond the health sector alone. Local governments, education systems, community organisations, technology developers, and the private sector all have a role to play. Cross-sectoral partnerships are needed to develop environments, services, and systems that empower people to care for their own health throughout the life course. Policy should incentivise innovation and co-production, ensuring self-care is not a matter of personal resilience alone but a collective responsibility enabled by system design.

## Study limitations

The principal limitations of this study are concerned with reliance on self-reported data that introduces the possibility of social desirability bias, and the cross-sectional design precludes causal inferences regarding the observed associations. it cannot establish causality. Thus, while the study captures a snapshot in time from representative cross section of British society and associations between demographic characteristics and self-care confidence, health literacy and professional support are identified, causal inferences cannot be made. Longitudinal research is needed to assess how these factors evolve and interact over time.

Although the sample size was large (N = 3,255) and included participants from across the UK, the survey relied on convenience and snowball sampling via online channels and professional networks. Despite the relatively balanced and comparable demographic profile of respondents, we acknowledge that this may have introduced selection bias, favouring individuals who are more health-literate, digitally connected or engaged in self-care conversations. Consequently, the findings may underrepresent perspectives from digitally excluded populations, socioeconomically disadvantaged groups, or individuals with lower educational attainment. This limitation is especially relevant when interpreting findings related to digital health access and health literacy.

Because the survey was administered online, individuals without reliable internet access, adequate digital literacy, or comfort using digital tools may have been excluded further limiting the representativeness of the sample and may skew findings related to the use of digital health information or engagement with online self-care resources. For reasons of pragmatism, the survey was administered in English only and this necessarily excluded non-English speakers or those with limited English proficiency. Given the importance of inclusivity in self-care research, future studies should consider multilingual administration to better reflect the UK’s diverse population.

Further, although healthcare professionals were included as a subgroup within the sample, the survey was not powered to compare subgroups with high precision. Additionally, the sample of HCPs may not fully represent the diversity of roles, settings and regions within the NHS, potentially limiting the generalisability of professional perspectives on barriers to self-care promotion.

While the survey draws on validated tools such as the HLS19-Q12 and underwent piloting for usability and content validity, full psychometric validation (e.g., factor analysis, test-retest reliability) was not conducted as part of this initial study. This will be the premise of our future work as this would help strengthen the tool’s measurement properties to support wider adoption and repeat use in longitudinal and cross-national studies. We also acknowledge that although regression models adjusted for key demographic factors, there remains the possibility of residual confounding by unmeasured variables such as income, occupation type, cultural beliefs, or personal health history, which could influence self-care behaviours and perceptions.

## Conclusion

This study advances our understanding of self-care confidence, professional guidance and health literacy within the UK population. The findings underscore the critical role of HCPs in fostering self-care engagement and highlight the need for targeted interventions to address health literacy deficits and structural barriers. Overall, the study findings paint a picture of a population that appears to understand the notion of self-care and is willing to engage but does not feel supported to do so. Paradoxically, professionals feel that public understanding and engagement are problematic, with health literacy gaps and personal understandings of self-care at the root of this apparent paradox.

Moving forward, a more inclusive, equitable approach to self-care promotion that rectifies incongruent understanding is necessary to ensure that all individuals, regardless of age, socioeconomic status or digital literacy, can actively participate in their health management. Future research should explore longitudinal impacts of self-care interventions and clarifications and the effectiveness of tailored health literacy programs in enhancing self-care autonomy.

## Data Availability

All relevant data are within the manuscript and its Supporting Information files.

## Declarations

### Data sharing statement

The data that support the findings of this study are contained within S3 File.

## Acknowledgments

The authors thank the Self-Care Forum for disseminating the link to the survey

## Author Contributors

All authors provided substantial contributions to the conception (PS, AEO), design (AEO, PS, SA, MA), acquisition (SA, AEO) and interpretation (PS, SA, MA, DM, RB, AEO) of study data and approved the final version of the paper. AEO took the lead in planning the study with support from co-authors. SA and MA carried out the data analysis with support from AEO. AEO is the guarantor.

**Twitter:** @austenelosta @ImperialSCARU

## Supporting Information

**S1 File: Survey export**

**S2 File: Survey findings**

**S3 File: Raw Data.xls**

**S1 Table: Main barriers to self-care for your patients/clients S2 Table: State of health, wellbeing and satisfaction.**

**S1 Fig: Factors affecting confidence caring for own health with guidance and support from HCP.** Forest plot showing the odds ratios and confidence intervals of factors associated with being more confident about caring for own health with guidance and support from an HCP.

**S2 Fig: Factors associated with not wanting more responsibility over own health.** Forest plot showing the odds ratios and confidence intervals of factors associated with not wanting more responsibility over own health.

**S3 Fig: Encouragement by HCP to play more active role in staying healthy** and **treating common conditions**. Forest plot showing the odds ratios and confidence intervals of factors associated with being encouraged by GP/nurse/pharmacist to play a more active role in staying healthy and treating common conditions by oneself.

